# Infection inhibiting effect of RT-PCR testing-isolation in COVID-19 - a case study of Hiroshima and Fukuoka in Japan -

**DOI:** 10.1101/2021.08.24.21262517

**Authors:** Kazuo Maki

## Abstract

A simple method of estimating the effect of reverse transcription polymerase chain reaction (RT-PCR) testing-isolation on the restraint of infection of COVID-19 is proposed. The effect is expressed as the ratio *χ* of the reproductive number to that in the case that no isolation measure would be taken. The method was applied in the case of the third infection wave (from December, 2020 to February, 2021) of Hiroshima and Fukuoka in Japan. The ratio *χ* was estimated to be 0.78 to 0.84 and 0.86 to 0.9 in Hiroshima and Fukuoka, respectively. It is also shown that the reduction of *χ* by 0.07 would have reduced at least 50% of total infected patients during the third infection wave in Fukuoka.

## 1. Introduction

COVID-19 has a unique characteristics that the primary infected patient causes secondary infection before development of symptom [1]. Isolation of infectious patients should be based on a reverse transcription polymerase chain reaction (RT-PCR) testing rather than on a symptom development. In order to consider a strategy to an infection measure, it will be useful to estimate how much the RT-PCR testing-isolation is effective in the infection restraint. This report proposes a simple method of that estimation, which is a generalization of the method described in the Appendix of Ref. [2].

The method is described in the next section, and it is applied in the case of Hiroshima and Fukuoka in Japan in section 3. The infection profile from December, 2020 to February, 2021 (the third infection wave in Japan) is analyzed in section 4. The last section is devoted to a conclusion and discussion.

## 2. Effect of RT-PCR testing-isolation on the restraint of infection

A direct index of the infection prevalence is the effective reproductive number *R*_t_, which means how many secondary infection is caused on average by a primary infected patient. If the reproductive number in the case that no test-isolation measure would be taken is denoted as *R*_t0_, the effect of RT-PCR testing-isolation can be expressed as a factor *χ* which is defined as follows.

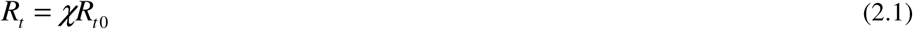

The factor *χ* means the ratio of secondary infection realized before the isolation of the primary infected patient to the whole infection without isolation. There are two kinds of time profile of secondary infection. One is generation time distribution, and the other is TOST (time from onset of symptom to transmission) distribution. (TOST distribution is called “infectiousness profile” by Xi He et al. [1].) It is known that TOST describes more accurately the timing of secondary infection [3].

Here, TOST distribution is expressed as *f*_i_, where *i* means the lapsed days from the onset of symptom. The accumulation *F*_i_ of *f*_j_ with regard to *j* up to *i*-1 means the expected value of proportion of infection completed by the timing *i* [3].

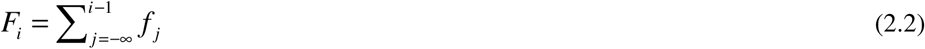

If all the infected patients are isolated at a timing *i* in the TOST distribution, the expected factor of the RT-PCR testing-isolation is given by *χ*=*F*_i_. Therefore, if the isolation timing *i* has a distribution of *p*_i_,

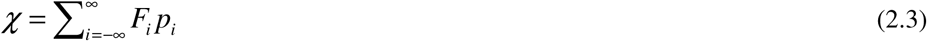

It should be noted that *p*_i_ does not represent the proportion of actually isolated patient at the timing *i*, but that *p*_i_ represents the proportion of patient tested at the timing *i*. Whether tested patient is isolated or not is determined by the test result. The isolation timing distribution is related to the distribution *ρ*_i_ of time (delay from onset of symptom to RT-PCR test-positive confirmation, which can be deduced from the data of symptom onset days. In this report, all the positively confirmed patients were assumed to be isolated according to the law applied to COVID-19 in Japan.

If the test timing is before the onset of symptom, RT-PCR test-positive probability is increasing. Since test-negative patients become positive later and that they are expected to be tested again, only the RT-PCR test-positive patients should be counted in the estimation of *p*. In this report, the time from RT-PCR test (swab sampling) to the confirmation of test result was assumed to be 1 day. Thus *p*_*i*_ ∝ *ρ*_*i*_, when *i*-1 < 0. On the other hand, if the test is done after the onset of symptom, RT-PCR test-positive probability is decreasing. In that period, since RT-PCR test-negative patients are not positive any more, all the tested patients should be counted in the estimation of *p*. Otherwise, patients who were test-positive but become test-negative at the test timing would not be counted as infected patients. Thus *p*_*i*_ ∝ *ρ*_*i*_ / *g*_*i*-1_, when _*i*-1_ ≥ 0. Here, RT-PCR test positive probability of infected patient at the test timing *i* is denoted as *g*_i_., which was estimated by a survival analysis [4]. The estimation in the case of upper respiratory tract sampling was used in this report.

In order to simplify the equation, the value of *g*_i_ is defined as 1 for *i* < 0. If the proportion of patients who do not have RT-PCR test is denoted as *p*_∞_, *p*_i_ is expressed as follows.

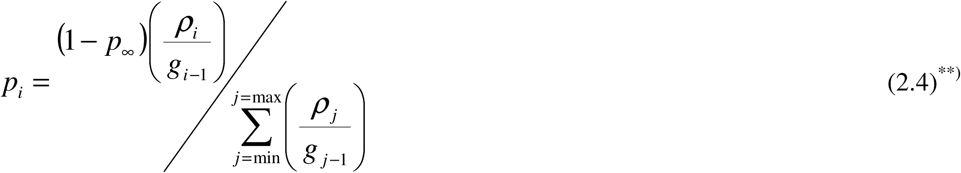

Here, min and max mean the time range of the distribution *ρ* and *g*. The ratio of *R*_t_ to *R*_t0_, *χ* is expressed as

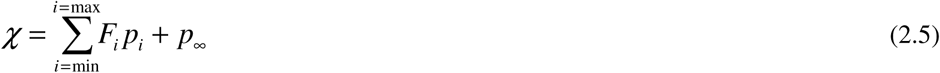

Since there is no reliable information on *p*_∞_, the contribution of *p*_∞_ was neglected in this report, but will be discussed in the last section.

In this report, distributions of incubation period, generation time, and TOST estimated by L. Ferretti et al. [3] were used, and RT-PCR test-positive probability profile estimated by S. Mallette et al. [4] was used. These are shown in Figs. 1, 2, and 3.

**Fig. 1.**
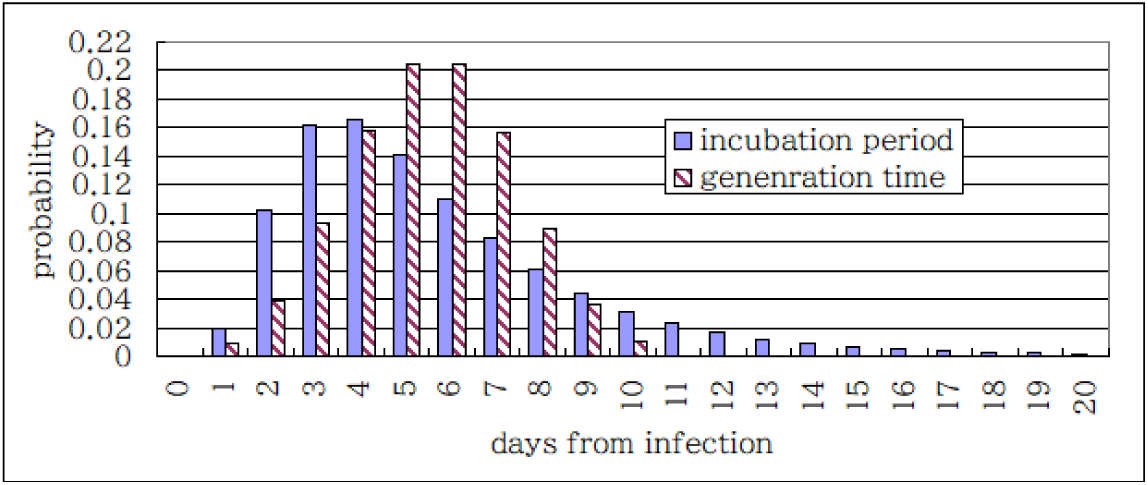
Incubation period and generation time distribution.

**Fig. 2.**
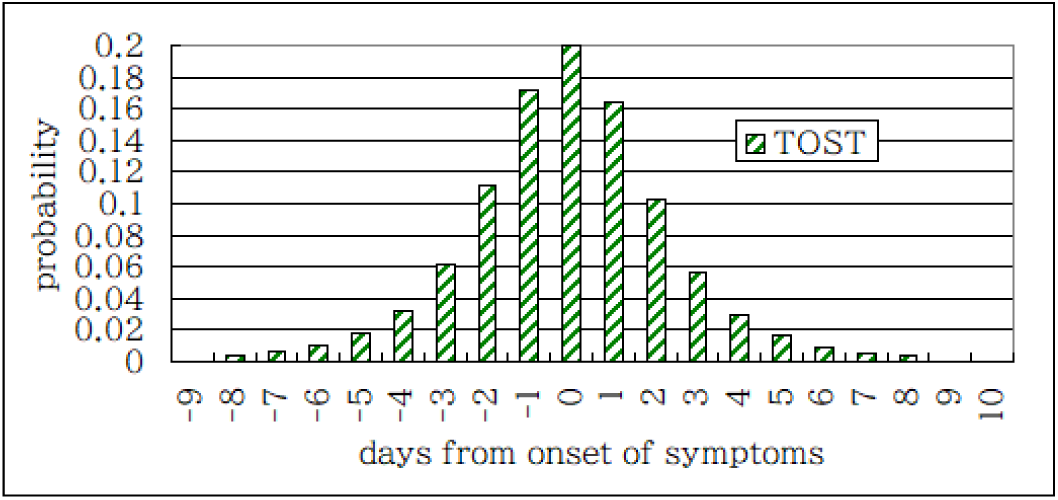
TOST (time from onset of symptom to transmission) distribution.

**Fig. 3.**
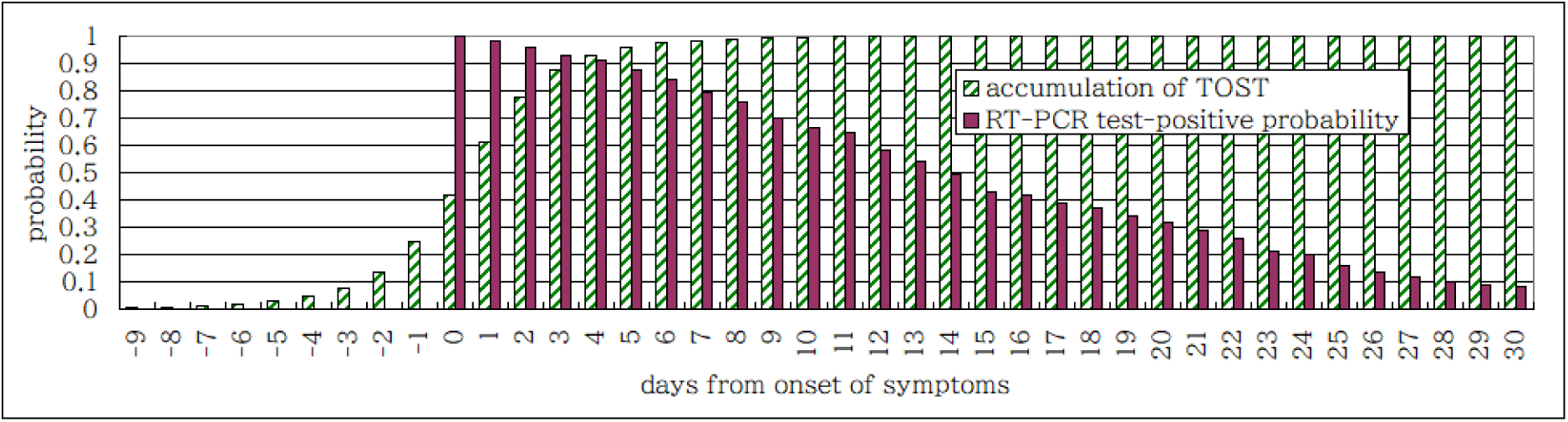
RT-PCR test-positive probability and the accumulation of TOST distribution.

## 3. Estimation of the effect of RT-PCR testing-isolation on the restraint of infection in Hiroshima and Fukuoka

Hiroshima and Fukuoka are typical cities in the west side of Japan. Population is similar to each other, about 1,200,000 and 1,600,000, respectively. Time (delay) from onset of symptom to PCR test-positive confirmation was picked up from the public data sources: Hiroshima city a [5], Hiroshima prefecture [6], Fukuoka city [7], and Fukuoka prefecture [8]. The interval is from October 15 in 2020 to March 3 in 2021. The delay distribution *ρ* from the available data is shown in Fig.4.

**Fig. 4.**
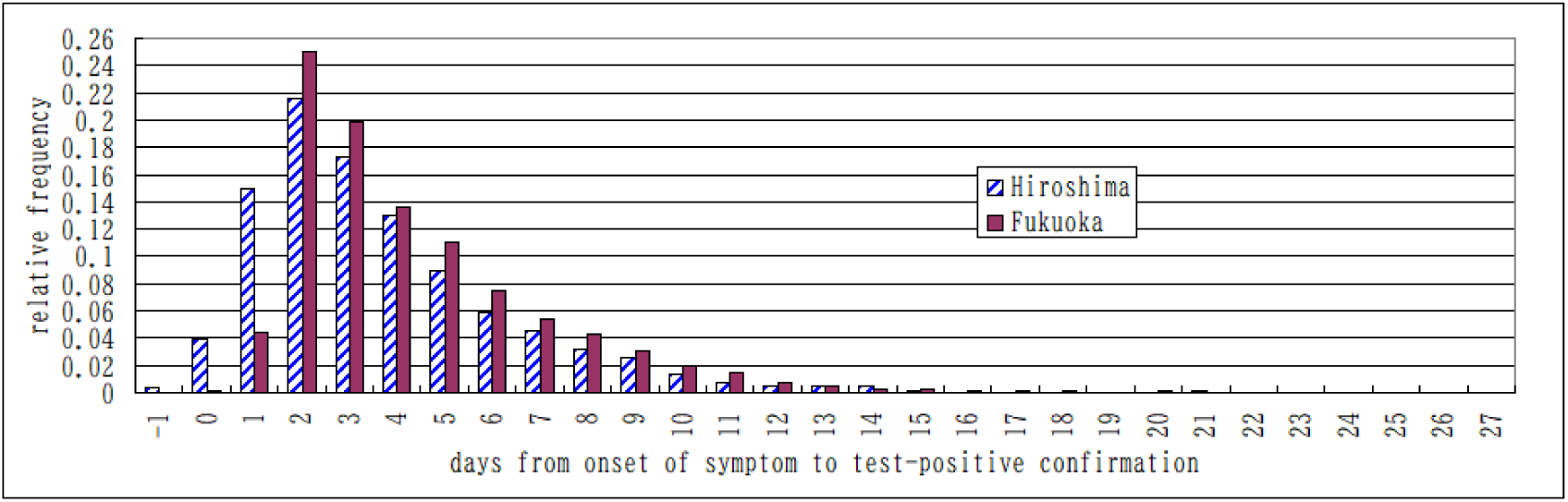
RT-PCR test-positive confirmation delay distribution of Hiroshima and Fukuoka.

The average delay from the onset of symptom to the confirmation of positivity is found to be 3.6 days and 4.3 days in Hiroshima and Fukuoka, respectively. Since swab sampling (testing) was assumed to be made one day before the test confirmation day, the positions -1 and 0 of the transverse axis in Fig.4 correspond to the symptom onset of 2 days and 1 day after the test confirmation day, respectively. These are interpreted as “presymptomatic” cases. Therefore, patients with known onset date of symptom were divided into two categories: A. Patients who had already developed symptom on the RT-PCR test day or did on that day; B. Patients who developed symptom after the RT-PCR test day. However, there are many cases that the date of the onset of symptom is not known. Such cases were divided into three categories: C. There had been onset of symptom, but the date is not known; D. There had been no onset of symptom until the test timing; E. It is not known whether there had been onset of symptom or not. The whole RT-PCR test-positive patients were divided into these five categories, and numbers of the patients belonging to these categories are summarized in Table 1. Hereafter, they are denoted as *n*(A), *n*(B), *n*(C), *n*(D), and *n*(E).

**Table 1.** Sorting of RT-PCR test-positive patients and estimation of presymptomatic cases.

| Category | onset date | symptom onset<br>on or before PCR test | symptom onset<br>after PCR test | Hiroshima | Fukuoka |
| --- | --- | --- | --- | --- | --- |
| A. | known | Yes | No | 2232 | 5574 |
| B. | known | No | Yes | 89 | 7 |
| C. | unknown | Yes | No | 1 | 17 |
| D. | unknown | No | unknown | 296 | 20 |
| E. | unknown | unknown | unknown | 310 | 601 |
| Total |  |  |  | 2928 | 6219 |
| NS | known or unknown | No | unknown | 385 to 695 | 27 to 628 |
| Proportion |  |  |  | 13.1 to 23.7% | 0.4 to 10.1% |
| PS (c.i.95%). | known or unknown | No | Yes | 149 to 404 | 19 to 342 |
| Proportion |  |  |  | 5.1 to 13.8% | 0.3 to 5.5% |
| AS (c.i.95%) | No | No | No | 159 to 418 | 20 to 355 |
| Proportion |  |  |  | 5.4 to 14.3% | 0.3 to 5.7% |

In order to estimate *χ*, it is necessary to estimate the symptom onset timing relative to RT-PCR test-confirmation day. Especially, it is important to estimate the number of presymptomatic patients. Patients who had no symptom onset before the RT-PCR test day is denoted as NS. The number of the patients NS (*n*(NS)) is not known, but the number is from *n*(B)+*n*(D) to *n*(B)+*n*(D)+*n*(E). In this report, the probability that *n*(NS)=*i* (denoted as *P*(NS)_i_) was assumed as follows.

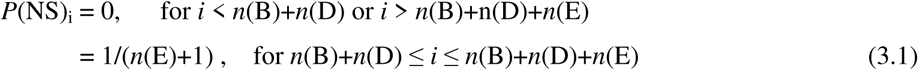

Patients in the category NS were further divided into two categories: PS: presymptomatic; AS: asymptomatic. The patients in the category PS develop symptom after the RT-PCR test day, and among those in PS, those in the category B have definite onset days. The proportion of presymptomatic infection (*x*) in the whole non-symptomatic patient at the RT-PCR test timing was evaluated by J. He et al. [9]. The estimated probability of the proportion *x* follows a normal distribution with average=0.4889 and standard deviation=0.08837, which is denoted as *norm*(*x*, 0.4889, 0.08837). Therefore, the probability that the total number of presymptomatic patients = *i* may be assumed as,

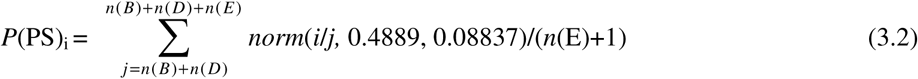

The lower and upper edges of the 95% confidence interval of the number of total presymptomatic patients are denoted as *n*(PS)^lower^ and *n*(PS)^upper^, respectively. The corresponding number of patients who have had symptoms before or on the day of the test without known symptom onset day (C and part of E with symptom) was estimated as an expectation on the condition that *n*(PS) is fixed. These numbers were used to calculate the confidence interval of *χ*.

The 95% confidence interval of presymptomatic patients without known symptom onset day is *n*(PS)^lower^ - *n*(B) to *n*(PS)^upper^ - *n*(B). The symptom onset day of these patients was assumed to be either 1 day or 2 days after the RT-PCR test day, which means that the confirmation delay distribution was assumed to be a delta function, either *δ*(*i*) or *δ*(*i*+1). This is an assumption to evaluate the RT-PCR testing-isolation effect and to compare two cities in the same condition. Since there are presymptomatic cases that the symptom onset day is more than 2 days after the RT-PCR test day [10], this evaluation may underestimate the effect of RT-PCR testing on the restraint of infection. The confirmation delay distribution of the symptomatic patients without known onset day of symptom (C and part of E with symptom) was assumed to be that of Fig. 4 without the presymptomatic part (−1 day and 0 day). Thus the final distribution *ρ*_i_ of time *i* from onset of symptom to RT-PCR test-positive confirmation was calculated as a weighted average of that of Fig. 4, that of Fig.4 without the presymptomatic part, and that of the δ distribution assumed for the presymptomatic patients without known onset timing.

It should be noted that this calculation focused only on the symptomatic patients including presymptomatic cases. The secondary infection by asymptomatic patients was assumed to have the same time profile as the symptomatic patients, and the peak of that time profile was assumed to play the roll of onset timing of symptom. The distribution of the delay from the peak of the secondary infection time profile to the confirmation of the asymptomatic patients was assumed to be the same as that of the final distribution as above.

The reduction factor *χ* of the reproductive number by RT-PCR test-isolation was calculated from Eqs. (2.4) and (2.5), and it is summarized in Table 2.

**Table 2.** Reduction factor *χ* of the reproductive number by RT-PCR test-isolation.

| Assumed symptom onset day of presymptomatic case | Hiroshima | Fukuoka |
| --- | --- | --- |
| 1 day after the test day | 0.80 to 0.84 | 0.87 to 0.89 |
| 2 days after the test day | 0.78 to 0.83 | 0.86 to 0.89 |

Table 2 shows that the RT-PCR test-isolation policy directly reduces the reproductive number by 10% to 30%. Although there remains some uncertainty, the difference between two cities is clear. The difference in *χ* is about 7%. There are two mutually correlated measures to be taken to reduce *χ*: (1) reducing the time (delay) from the onset of symptom to RT-PCR testing, and (2) testing and finding patients without symptom.

## 4. Analysis of the third infection wave in Hiroshima and Fukuoka

In this section, it is described how the analysis in the precedent section is related to the real situation. The infection profile from December, 2020 to February, 2021 is analyzed in Fig. 5. The number of daily confirmed RT-PCR positive patients is plotted as the blue rhombic mark. The number of patients who develop symptom is plotted as the red square mark. Here, as an approximation, the time profile of the effective symptom onset of the patients of the categories C, D, and E was estimated by a sum of the confirmation time sequence weighted by the confirmation delay distribution in Fig.4^***)^. The daily expectation number of newly infected patients was estimated by a back propagation method [11], [12] from the symptom onset profile and the incubation period distribution in Fig.1^****)^, and it is plotted as the yellow triangle mark. The reproductive number *R*_t_ is shown by the solid black line in Fig.5. It is given by the ratio of the number of new infections generated at time step *t* to the sum of infection incidence up to time step *t*-1 weighted by the generation time distribution [13]. The boundaries of 95% confidence interval of *R*_t_ are shown by two dotted lines.

**Fig. 5.**
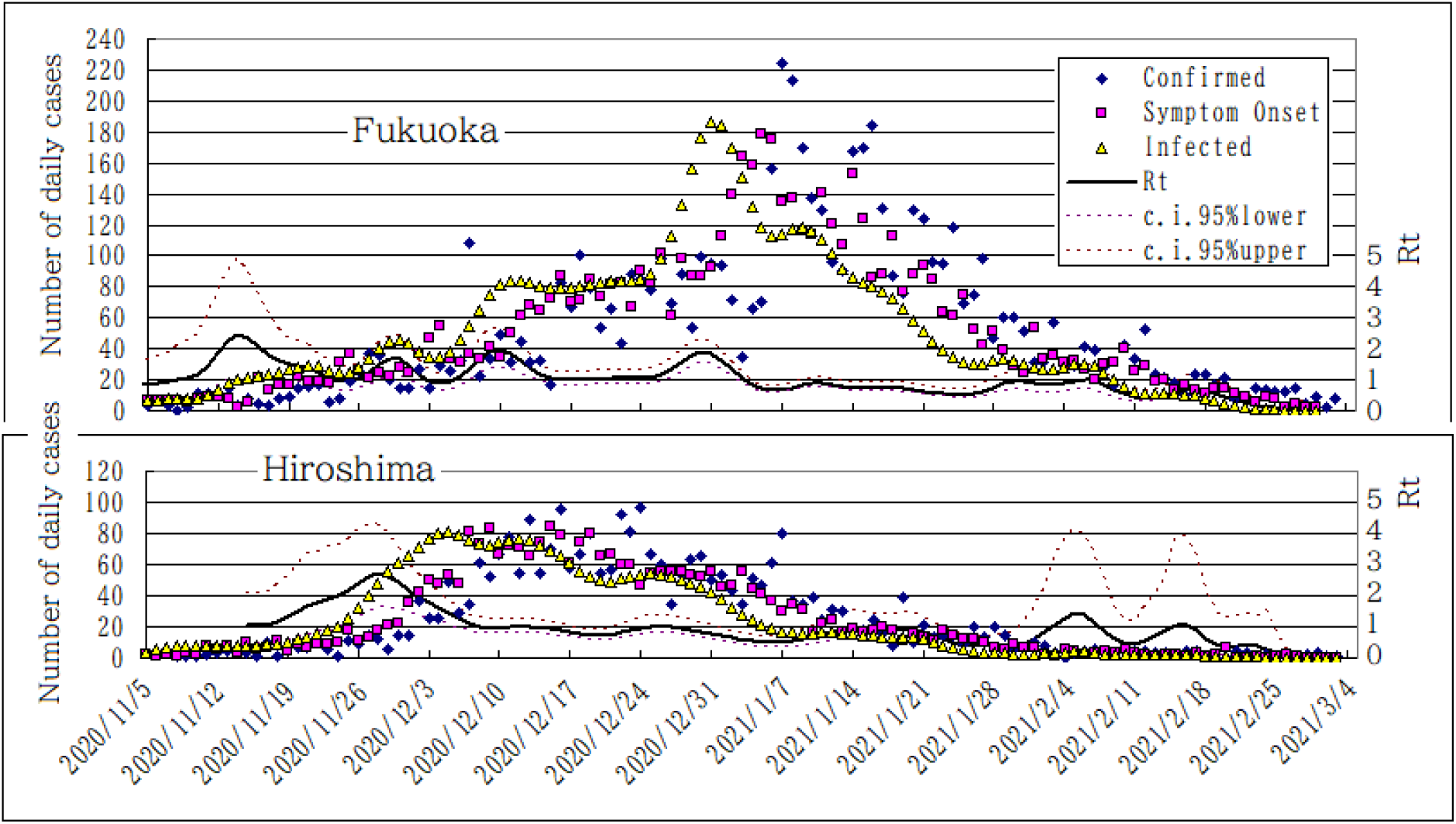
Infection profile of Hiroshima and Fukuoka.

There is an important difference between the two cities in December. The reproductive number *R*_t_ in Fukuoka is enhanced to around 2, while that in Hiroshima stays around 1. In every end of the year, people in Tokyo area and Osaka area tend to return to their homeland cities, and enjoy reunion with relatives. The rise in *R*_t_ in Fukuoka is thought to have been caused by this activities. However, in Hiroshima an infection prevention measure was put into effect from December 17, 2020 to January 3, 2021 (Hiroshima city b [14]). If a similar measure would have been taken in Fukuoka, *R*_t_ might have been reduced to the level of *R*_t_ of Hiroshima divided by 0.93 in the same interval. Time profile of infection in that case was calculated using the same generation time distribution, and it is shown as the white square mark in Fig.6. On the other hand, if Fukuoka would have carried out continuously the same level of RT-PCR testing-isolation as that of Hiroshima, *R*_t_ of Fukuoka would have been multiplied by 0.93. That case is shown as the red rhombic mark in Fig.6. This means that either the short term measure of social contact restriction or the long term measure of enhancement of RT-PCR testing would have restrained infection to the same level as that of Hiroshima.

**Fig.6.**
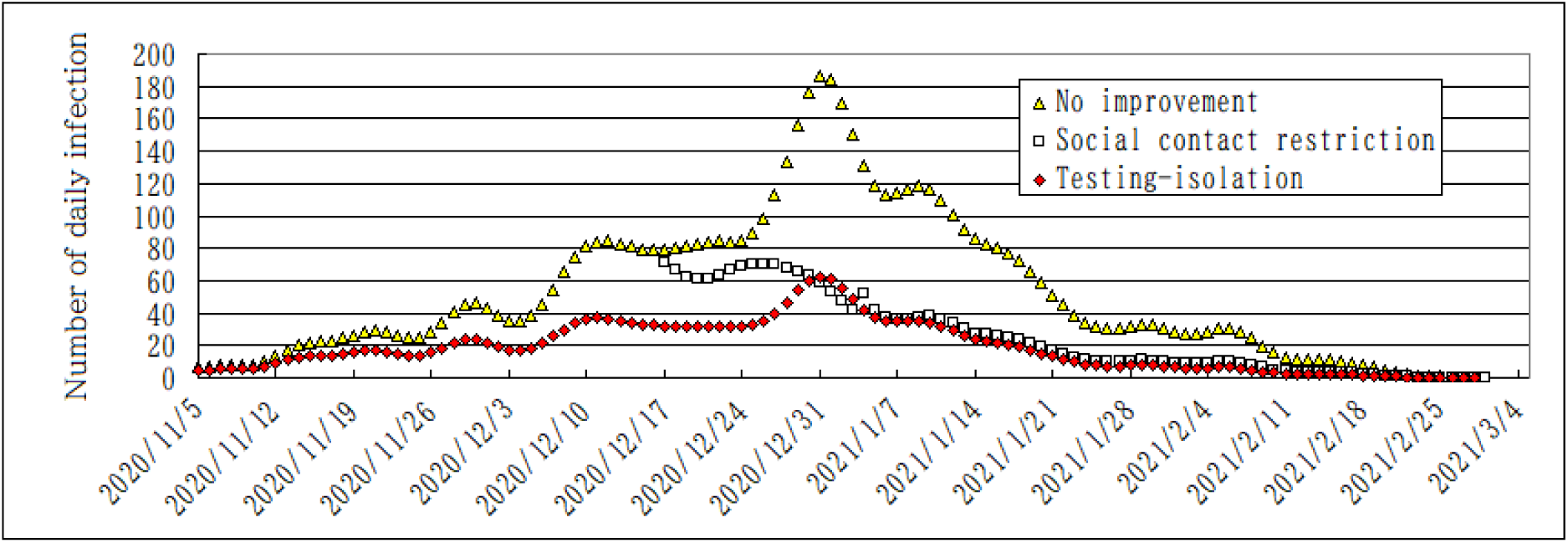
The expected effect of two measures to be taken in Fukuoka.

## 5. Conclusion and discussion

RT-PCR testing is essential to cope with COVID-19. The testing is necessary to starting treatment of patient. The test data is also important for the policy person in charge who takes infection counter measures. This report focuses on another roll of the RT-PCR testing: a direct contribution to infection restraint. If RT-PCR test-positive patients are appropriately isolated, the direct contribution can be estimated using the record of the symptom onset days. The method is very simple, but there remain uncertainties because of unknown records on symptom onset.

It was shown that the RT-PCR testing-isolation is directly reducing the reproductive number by 10% to 30% in Hiroshima and Fukuoka. The difference between these cities was clear and was about 7% on average. There are two measures to be taken to reduce the reproductive number by RT-PCR testing-isolation: (1) reducing the time from the onset of symptom to PCR testing, and (2) testing and finding patients without symptom.

The infection profile from December, 2020 to February, 2021 was analyzed. It was shown that either the short term measure of social contact restriction or the long term measure of enhancement of RT-PCR testing would have restrained the infection of Fukuoka to the same level as that of Hiroshima.

Some points should be noted regarding the applicability of the method proposed in this report.

a. This method is based on the fact that every RT-PCR test-positive patient is appropriately isolated. Otherwise, this method would overestimate the effect.
b. Time from onset of symptom to transmission (TOST) distribution is estimated from the records of many infector-infectee transmission pairs. These data are thought to be affected by RT-PCR testing-isolation. The actual distribution without isolation may have a longer tail than that applied here. If that is the case, this report may underestimate the effect.
c. This method requires a symptom onset day of every patient. The result is dependent on how to assign an onset day to the patient without known symptom onset day. However, a comparative study may not be affected much, if the assignment policy is determined consistently.
d. This method neglects the effect of patients who are not tested at all. Most part of such patients is thought to be asymptomatic. If the proportion (*p*_∞_) of such patients is known, the effect can be estimated as Eqs. (2.4) and (2.5). The 95% confidence interval of the number of asymptomatic patients tested-positive (AS) may be estimated by Eq. (3.2) by replacing 0.48889 with 0.51111. The proportion among the total test-positive patient is shown in the last line in Table 1. This proportion is denoted as *a*. On the other hand, the proportion of asymptomatic patient among all the infected patients is estimated to be between 9% and 26% in many meta-analysis papers [9], [15], [16]. This proportion is denoted as *b*. The difference between these proportions comes mostly from the asymptomatic patients who are not tested at all. If all the undetected patients are asymptomatic, the following relation holds.

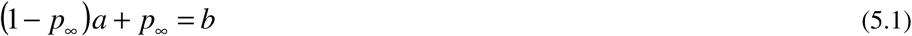

If *b*=0.20, then *p*_∞_ is estimated to be from 0.067 to 0.154 and from 0.152 to 0.198 for Hiroshima and Fukuoka, respectively. In order to decrease this proportion (*p*_∞_), it is necessary to expand further more the target of RT-PCR testing.

## Data Availability

For this study, I used publicly available data of COVID-19 provided by Hiroshima city, Hiroshima prefecture, Fukukoka city, and Fukuoka prefecture in Japan. The data links are given in the manuscript.

https://www.city.hiroshima.lg.jp/site/korona/108656.html

https://www.pref.hiroshima.lg.jp/soshiki/19/opendata-covid19.html

https://www.city.fukuoka.lg.jp/hofuku/hokenyobo/health/kansen/cohs.html

https://www.pref.fukuoka.lg.jp/contents/covid19-hassei.html

## Footnotes

**) It should be noted that the denominator means the expected ratio of the number of infected and tested patients to that of test-positive patients. This number was found to be from 1.25 to 1.28 in the case study of Sec. 3.

***) Details of the delay time profile discussed in Sec. 3 do not affect much the result of this kind of analysis to estimate *R*_t_.

****) The initial trial solution was given by a sum of the number of the symptom onset patients weighted by the incubation period distribution. The estimation-maximization step was iterated 15 times. In each step the solution was smoothed by a binomial filter with window width of 5 days.

## Notes

### Competing Interest Statement

The authors have declared no competing interest.

### Clinical Trial

There is no clinical trial.

### Funding Statement

This research did not receive any specific grant from funding agencies in the public, commercial or not-for-profit sectors.

### Summary of Updates

The epidemic data of Hiroshima-city and Fukuoka-city are uploaded. The data of Figures 1, 2, and 3 are uploaded. The manuscript is not revised.

